# Association between PM_10_ exposure and risk of myocardial infarction in adults: a systematic review and meta-analysis

**DOI:** 10.1101/2023.07.21.23292792

**Authors:** Kleiton Strobl, Syed Asad Irfan, Hassan Masood, Noor Latif, Om Kurmi

## Abstract

**Background:** Air pollution has several negative health effects. Particulate matter (PM) is a pollutant that is often linked to health adversities. PM_2.5_ (PM with an aerodynamic diameter of ≤2.5μm) exposure has been associated with negative cardiovascular (CV) outcomes. However, the impact of PM_10_ (PM with an aerodynamic diameter of ≤10μm) exposure is often overlooked due to its limited ability to pass the alveolar barrier. This study aims to assess the association between PM_10_ exposure and risk of myocardial infarction (MI) amongst adults (≥18 years of age) as this has been poorly studied.

**Methods:** The study protocol was published on PROSPERO (CRD42023409796) on March 31, 2023. Literature searches were conducted on 4 databases (OVID Medline, Embase, CINAHL, and Web of Science) for studies looking at associations between PM and MI. English studies from all time periods were assessed. Studies selected for review were time-series, case-crossover, and cohort studies which investigated the risk of MI as an outcome upon PM_10_ exposure. The quality of evidence was assessed using Cochrane’s GRADE approach. Data for different risk outcomes (risk ratio (RR), odds ratio (OR), hazard ratio (HR)) and 3 lags was meta-analyzed using an inverse variance statistical analysis using a random effects model. The pooled effect sizes and the 95% confidence intervals (CIs) were reported in forest plots.

**Results:** Among the 1,099 studies identified, 41 were included for review and 23 were deemed eligible for meta-analysis. Our analysis revealed that there is an increased risk (OR=1.01; 95% CI:1.00 - 1.02) of MI with a 10 μg/m^3^ increase in PM_10_ after a lag 0 and lag 1 delay.

**Conclusions:** Our findings indicate that PM_10_ exposure is associated with an increased risk of MI. This can aid in informing environmental policy-making, personal-level preventative measures, and global public health action.

## 1. Introduction

Air pollution is a complex combination of gaseous and particle constituents, which are hazardous to human health [1]. In air pollution, particulate matter (PM) comprises carbonaceous particles containing adsorbed organic compounds and reactive metals [1]. Nitrates, sulfates, polycyclic aromatic hydrocarbons, endotoxin, and metals such as iron, copper, nickel, zinc, and vanadium are all common constituents of PM [1]. PM can be further classified relative to the particle size into coarse (PM_10_: diameter <10μm), fine (PM_2.5_: diameter <2.5μm), and ultrafine (PM_0.1_: diameter <0.1μm) [1]. Earlier scientific research has reported that PM_2.5_ is associated with adverse health outcomes, including poor cardiovascular (CV) health outcomes. PM_10_ is present in dust from roads, farms, construction sites, and mines [2]. However, the chemical composition and size distribution of PM_10_ varies widely depending on where it originates and how it forms in the environment [3]. Although PM_10_ is also associated with adverse impact health as it is an irritant for the nose, throat, and eyes; however, in general, this is not a major focus airborne pollutant of study [2].

The short-term impacts of PM_10_ exposure on respiratory pathologies, such as chronic obstructive pulmonary disease and asthma, are well known and studied [3]. However, in CV health research, the effects of PM_2.5_ are more often researched as PM_2.5_ has been shown to pass the alveolar barrier and cause an inflammatory response in blood vessels [4]. This exacerbates atherosclerosis and increases the risk of myocardial infarction (MI), ischemic heart disease, and thrombotic stroke [4]. The risk of MI was chosen as it is one of the largest causes of death worldwide, with approximately three million deaths annually [4]. Different populations are also exposed to varying levels of PM, and understanding its health adversities is important for understanding subsequent health disparities. Interestingly, there is a lack of up-to-date research regarding the impact of PM_10_ on the risk of MI despite instances being reported where exposure to PM_10_ has led to a surge in CV-related hospital admissions [3]. Although there is a higher probability of PM_2.5_ passing the alveolar barrier, it is still possible for PM_10_ to enter the bloodstream in smaller proportions [5]. When searching this topic on the PROSPERO database, only one matching systematic review and meta-analysis was found, and it solely assessed the impact of PM_2.5_ and not PM_10_ [6]. Past studies looked at MI risk due to short-term PM_10_ exposure, or long-term PM_10_ exposure, or both but are missing up-to-date literature in their review [7–9]. The primary aim of this systematic review and meta-analysis is to assess the effect of PM_10_ on the risk of MI to better understand its burden of disease and update pre-existing literature looking at the CV impacts of PM exposure.

## 2. Methods

### 2.1. Search strategy

Studies were obtained from a systematic search of the following databases: OVID Medline, Embase, CINAHL, and Web of Science. The literature searches were conducted on Jan 17, 2023, and included studies from all time periods. See the specific search terms utilized (S1 Table). The study protocol was published on PROSPERO (CRD42023409796) on March 31, 2023, after completing the literature search.

### 2.2. Inclusion/exclusion criteria

The inclusion criteria for this study were: (1) study design had to be time-series, case-crossover, or cohort; (2) PM_10_ had to be an exposure; (3) risk of myocardial infarction had to be an outcome (risk ratio/relative risk (RR), odds ratio (OR), hazard ratio (HR)); (4) population ≥18 years of age. Cohort studies were included after the protocol to increase the data pool.

The exclusion criteria for this study were: (1) Other primary study designs (RCTs, clinical trials, cross-sectional designs, protocols, pilot studies, etc.); (2) secondary studies (narrative reviews, systematic reviews, meta-analyses, scoping reviews); (3) Risk outcome(s) only reported in graph(s) (4) studies that only look at PM_2.5_ or coarse particulate matter (PM_2.5-10_); (5) population <18 years of age.

### 2.3. Lag periods

MI risk was stratified by exposure duration and lags were developed for the following reasons: (1) preservation of data validity and reliability and (2) lack of standardized short-term and long-term parameters for PM exposure. PM_10_ exposure lags were based on the following definitions: lag 0 = same day (0-24 hours); lag 1 = 1-3 day delay (24-96 hours); lag 2 = 3 day delay or more (>96 hours). If there were multiple data points, the value closest to the midpoint of the range was selected for meta-analysis for lag 0 and 1 (lag 0 = 12 hours; lag 1 = 60 hours). For lag 2, the value closest to >96 hours was selected for meta-analysis.

### 2.4. Study selection

Covidence was used to manage the screening phase of this study [10]. For the abstract screening, two authors (HM & NL) independently screened abstracts based on the inclusion/exclusion criteria, and another two (KS & SI) resolved conflicts. For full-text screening, two authors independently (KS & SI) screened the full manuscripts, and the same authors (KS & SI) discussed any conflicts and reached a consensus. The study drafting process was recorded using “Preferred Reporting Items for Systematic Reviews and Meta-Analyses” (PRISMA) guidelines [11].

### 2.5. Data extraction

Two authors independently extracted data from one half of the included studies (KS & SI), while another two (HM & NL) independently extracted data from the other half of the included studies. Any discrepancies in data values were corrected cohesively amongst the two authors for their extracted data half, respectively.

Data extracted from studies included: study name, design, country, sample size, male/female ratio, participant characteristics, exposure increments for risk measure, risk scale, lag intervals, and effect size (risk outcomes).

For studies with multiple models, outcome data values adjusting for the greatest number of confounding variables were meta-analyzed to maintain data validity. Industrial values were used for studies reporting industrial and non-industrial location outcome values because they were more likely to report reliable PM_10_ exposure MI risk outcomes. For studies that reported data on multiple regions of a country, data was selected from the most central geographical region.

### 2.6. Methodological risk of bias assessment

Risk of bias was assessed by two authors (KS & SI) using the NHLBI Quality Assessment Tool for Observational Cohort and Cross-Sectional Studies [12] which assesses methodological risk of bias through 14 criteria: (1) clear research question/objective; (2) clearly defined study population; (3) 50% participation rate of eligible persons; (4) all participants selected from identical/similar populations in the same time period using pre-specified inclusion and exclusion criteria; (5) provision of sample size justification, power description, or variance and effect estimates; (6) exposure measured before outcome in analysis; (7) sufficient time frame to expect association; (8) examination of differing levels of exposure as related to the outcome; (9) clearly defined, valid, reliable, widely-applied exposure measures; (10) >1 exposure assessments; (11) clearly defined, valid, reliable, widely-applied outcome measures; (12) blinding of outcome assessors to participant exposure status; (13) <20% loss to follow-up after baseline; (14) measurement and adjustment of potential confounders affecting impact between exposure and outcome.

Each criterion was deemed to be satisfied or unsatisfied based on the author’s rating. Overall quality ratings included: ‘good’, ‘fair’, and ‘poor’. The overall quality of each study began at ‘good’, and got demoted by one level per unsatisfied criteria. Conflicts in the methodological risk of bias assessment were resolved by the same two authors (KS & SI) through consensus.

### 2.7. Meta-analysis

Meta-analysis was conducted using Review Manager 5.4.1.

#### 2.7.1. MI risk outcome measures

The primary outcome was MI risk. For the meta-analysis, data was included for studies reporting risk based on PM_10_ concentration increments of 10µg/m^3^. MI risk outcome measures included RR, OR, and HR with 95% confidence intervals (CIs). A RR, OR, or HR >1 indicates a higher PM_10_-associated MI risk.

MI risk was stratified by MI risk outcome measure (RR, OR, HR) for the following two reasons: (1) to preserve data validity and (2) inherent distinctiveness of the outcome measures [13]. Data was also stratified by lags (0, 1, 2) as aforementioned.

#### 2.7.2. Data synthesis

The pooled risk outcome was considered if the following criteria were met: two or more studies reported the same MI risk outcome measure and lag, the I^2^ statistic for measuring statistical heterogeneity ≤60%.

An I^2^ >60% was considered heterogeneous on a statistically significant level. An inverse variance statistical analysis was conducted with a random-effects model when creating the forest plots, as there was heterogeneity, differences in study design, setting, and adjustment models. A P-value <0.05 was considered statistically significant.

Publication bias was assessed through analysis of funnel plot symmetricity. Publication bias was not assessed for quantitative analyses with <5 studies due to a lack of statistical power.

#### 2.7.3. Quality of evidence evaluation

Quality of evidence was evaluated by two authors (KS & SI) independently using Cochrane’s GRADE approach (Schünemann et al., 2013), which assesses: (1) risk of bias; (2) inconsistency; (3) indirectness; (4) imprecision; (5) publication bias; (6) large magnitude of effect; (7) dose-response gradient; (8) residual confounding. Overall quality ratings include: ‘high’, ‘moderate’, ‘low’ and ‘very low’. All resulting outcomes began with a rating of ‘high’ and were demoted one level for each unsatisfied criteria 1-5; upgrading could occur for satisfying criteria 6-8. Any evaluation conflicts were resolved by the same two authors (KS & SI) through consensus.

## 3. Results

### 3.1. Study selection

A breakdown of study identification and screening can be found in the PRISMA flowchart (Figure 1). A systematic literature search from databases/registers identified 1,099 potential studies viable for inclusion. After 396 duplicates were removed, 703 total studies were available for abstract screening. Five hundred sixty-nine studies were excluded following abstract screening, and 12 studies were not retrieved due to a lack of full-text availability. One hundred twenty-two studies remained for full-text screening, and 81 studies were excluded after the full-text screening: 64 for having the wrong outcome measure, 11 for having different study design (not meeting the inclusion criteria), 3 for analyzing PM_2.5_ exclusively and not PM_10_, 2 for having different patient population, and 1 study for not being in English. Forty-one studies were included for review, with 23 of 41 deemed eligible for meta-analysis.

**Figure 1.**
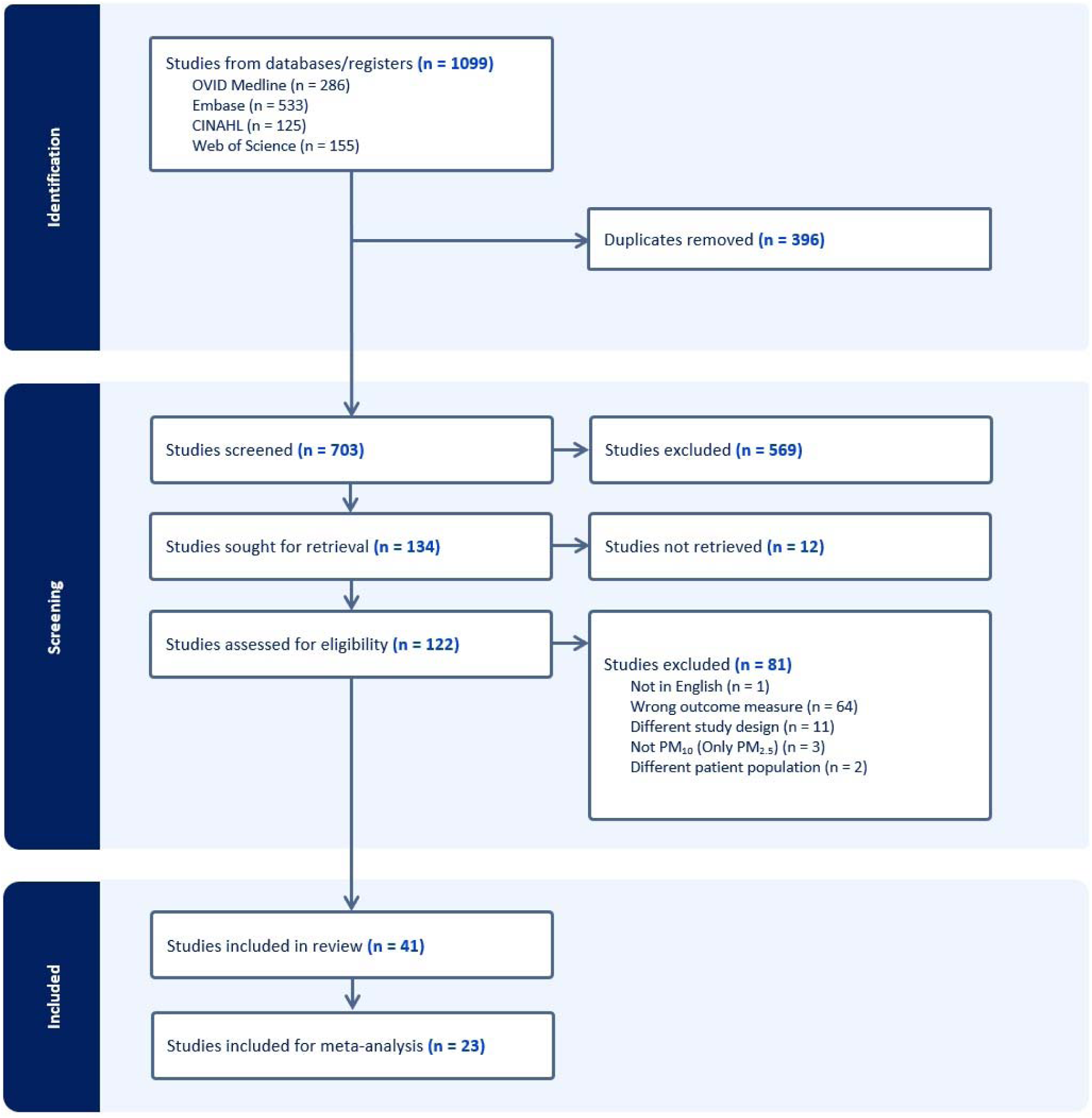
PRISMA flow diagram.

### 3.2. Study Characteristics

Table 1 provides the characteristics of studies included in the meta-analysis [14–36], adjusted confounding variables can be found for these respective studies (S2 Table). See study characteristics for other studies included in the review (S3 Table) (reference).

**Table 1.**
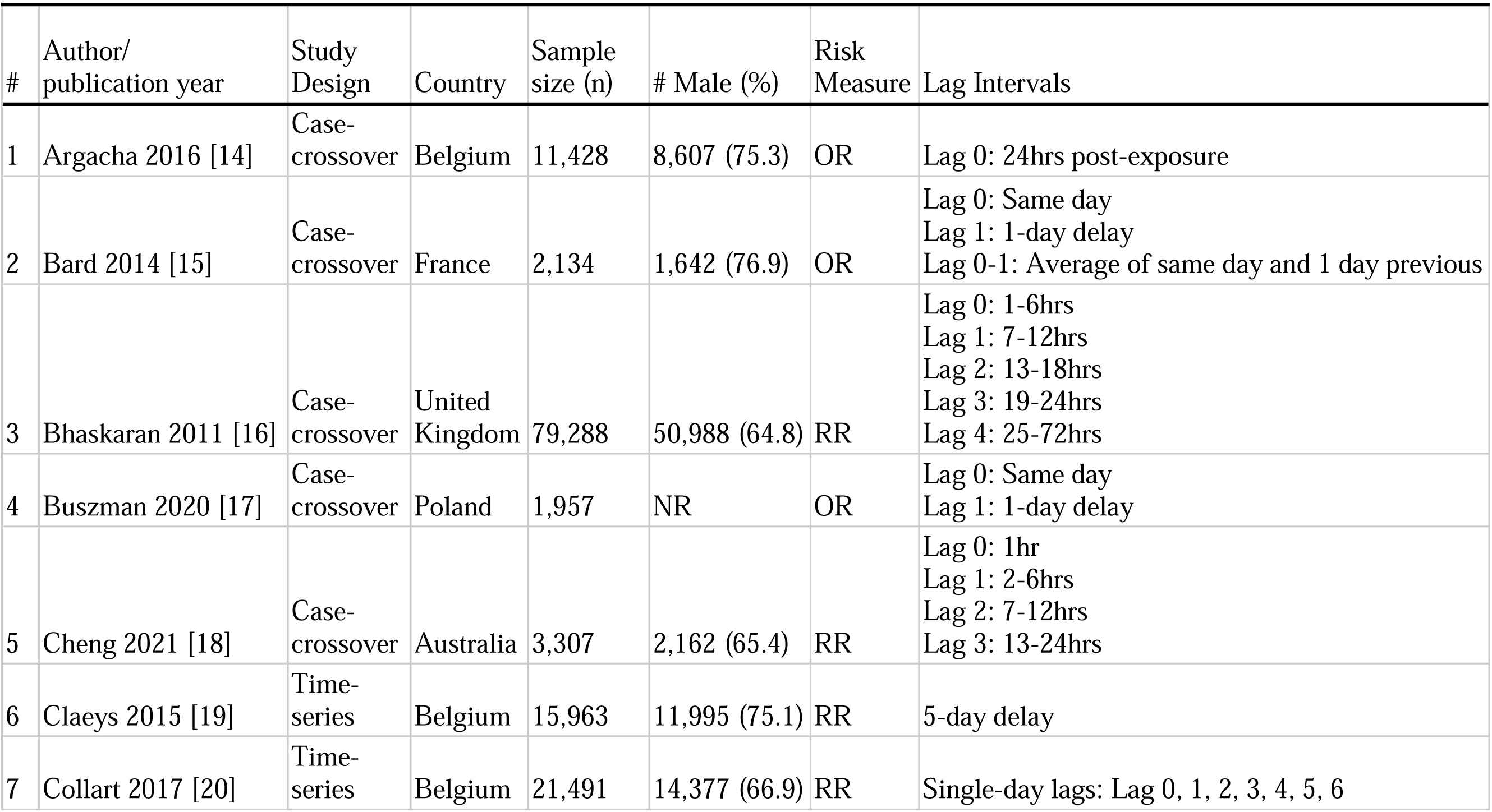
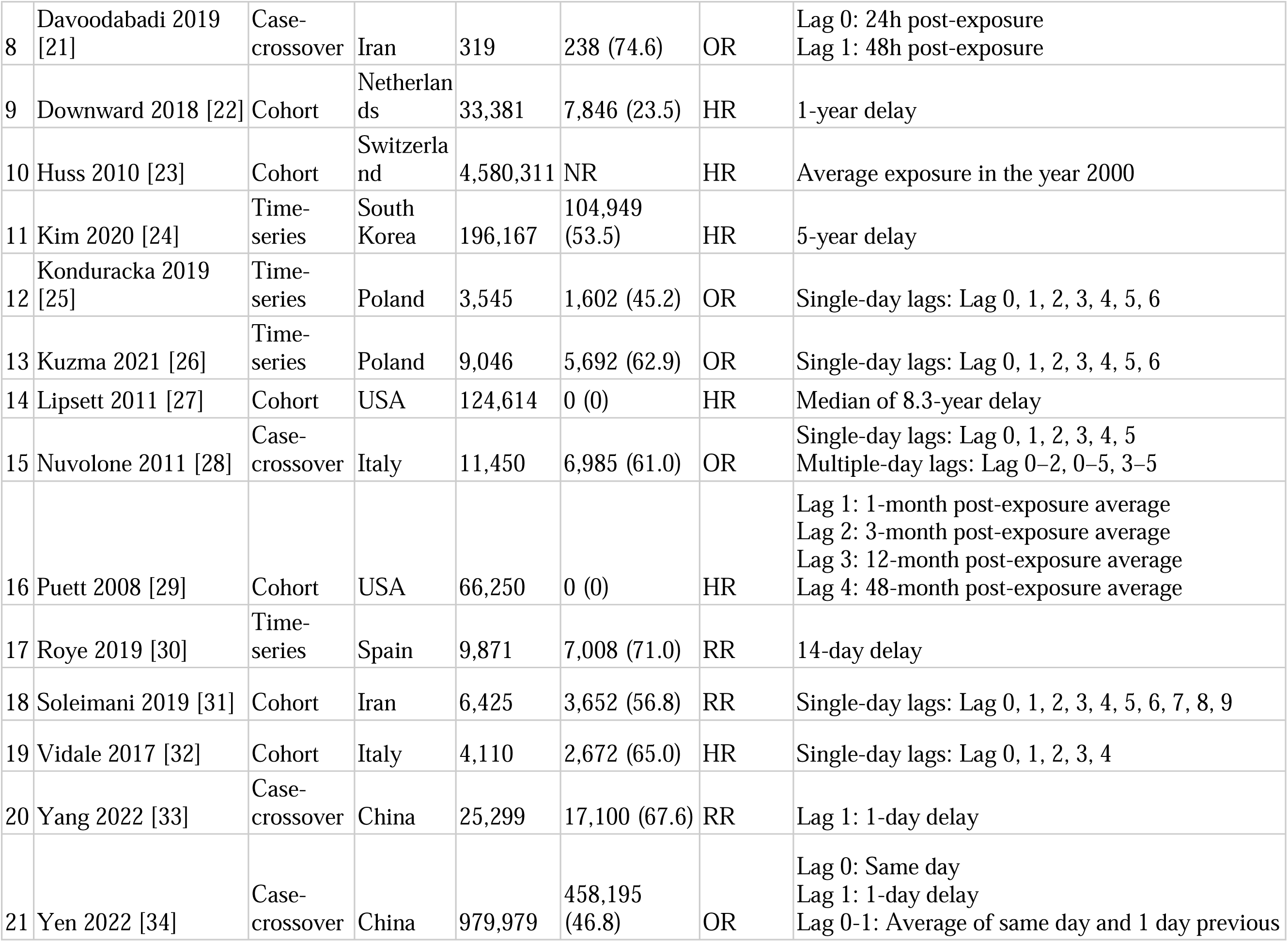
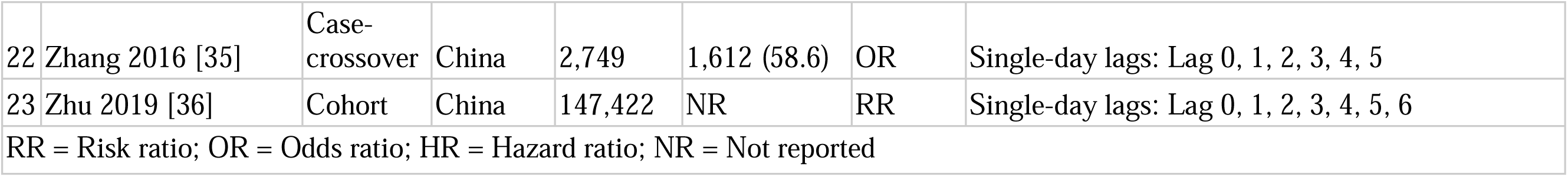
Study characteristics for studies included in the meta-analysis.

#### 3.2.1. Study design characteristics

Amongst the 41 studies included in the systematic review, 22 studies (53.7%) had a case-crossover design, 12 studies (29.3%) had an observational cohort design, and 7 studies (17.1%) had a time-series design.

For the meta-analysis, ten studies (43.5%) had a case-crossover design, seven studies (30.4%) had an observational cohort design, and six studies (26.1%) had a time-series design.

#### 3.2.2. Participant characteristics

For qualitative synthesis, data was calculated from a total of 6,663,870 participants. For the meta-analysis, data was calculated from a total of 6,336,506 participants. All but three studies reported a male/female participant ratio [17, 23, 36]. Of 1,606,816 participants from the remaining studies with a reported male/female participant ratio, there were 707,322 male participants (44.0%) and 899,494 (56.0%) female participants.

#### 3.2.3. MI risk outcome measures

Amongst the 41 studies included in the review, 18 studies (43.9%) used OR as their effect size for MI risk outcome measure, 12 studies (29.3%) used RR, and 11 studies (26.8%) used HR.

For the meta-analysis, studies most often utilized OR (n=9; 39.1%), followed by RR (n=8; 34.8%) and HR (n=6; 26.1%) for MI risk outcome measures.

### 3.3. Methodological risk of bias analysis for included studies

After the quality assessment was conducted (S4 Table), a large majority of studies were rated at a quality score of good (38/41), with the remainder of the studies rated at a quality of fair (3/41). No studies were rated at a poor quality score. All three studies demoted to a fair quality score were done so for the same reason: failure to adjust for confounders [17, 19, 42].

### 3.4. Primary MI risk outcomes – meta-analysis

See the meta-analysis summary of primary MI risk outcomes (Table 2). For lag 0 and lag 1 HR outcomes, meta-analysis could not be conducted as there was only one associated study for each respective outcome [32] (S1, S2 Fig). Evidence assessment could also not be conducted due to one study (S11, S12 Table).

**Table 2.**
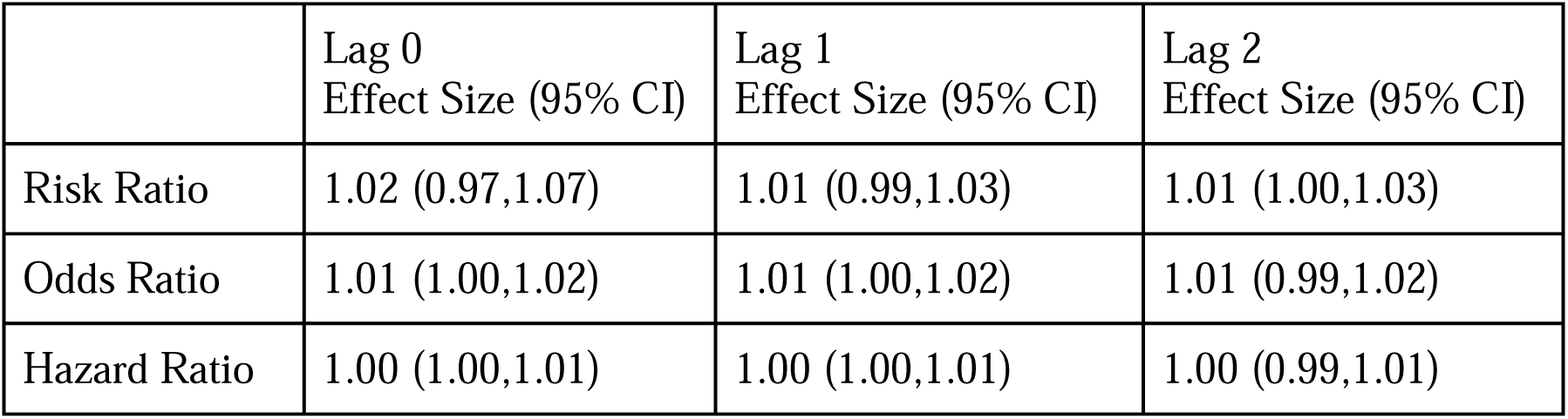
Summary of primary MI risk outcomes.

#### 3.4.1. Risk Ratio – Lag 0 (Same day [0-24hrs])

Three studies with three outcomes (n = 89,020) were reported for RR. Soleimani et al. [31] and Cheng et al. [18] reported an increased risk of MI with PM_10_ exposure, while Bhaskaran et al. [16] reported a decreased risk. Statistical pooling was appropriate due to statistical homogeneity (I^2^ = 60%) (Figure 2). Although there was an overall increased risk of MI (RR = 1.02, 95% CI: 0.97-1.07) it was not statistically significant (P-value = 0.43). Evidence quality was rated high on the GRADE scale (S5 Table).

**Figure 2.**
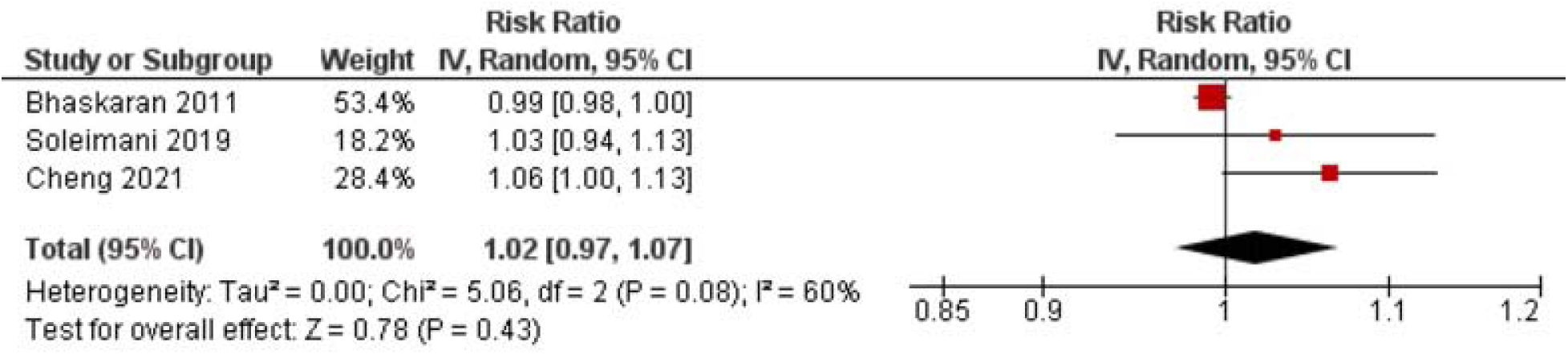
Lag 0 RR for MI after 10 μg/m^3^ increase in PM_10_ exposure.

#### 3.4.2. Risk Ratio – Lag 1 (1-3 days [24-96hrs])

Four studies with four outcomes (n = 132,503) were reported for RR. Statistical pooling was inappropriate for the lag 1 RR values because of statistical heterogeneity (I^2^ = 72%) (Figure 3). Evidence quality was rated moderate on the GRADE scale due to inconsistency (S6 Table).

**Figure 3.**
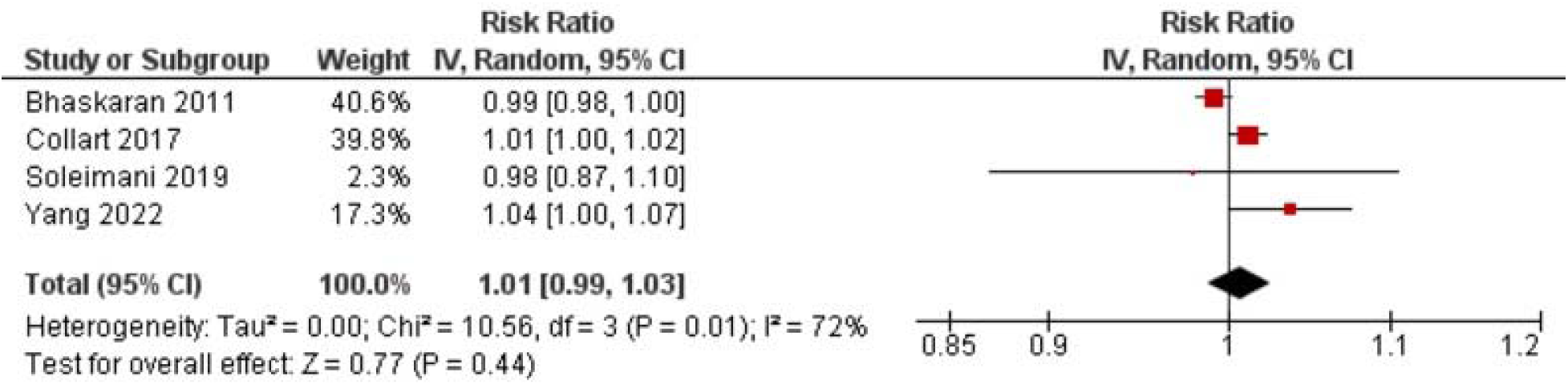
Lag 1 RR for MI after 10 μg/m^3^ increase in PM_10_ exposure.

#### 3.4.3. Risk Ratio – Lag 2 (3+ days [96hrs+])

Four studies with four outcomes (n = 179,681) were reported for RR. Roye et al. [30] and Zhu et al. [36] reported an increased risk of MI with PM_10_ exposure, while Soleimani et al. [31] reported a decreased risk. Claeys et al. [19] reported no change in risk. Statistical pooling was appropriate due to statistical homogeneity (I^2^ = 60%) (Figure 3). Although there was an overall increased risk of MI (RR = 1.01, 95% CI: 1.00-1.03), it was not statistically significant (P-value = 0.10). Evidence quality was rated high on the GRADE scale (S7 Table).

#### 3.4.4. Odds Ratio – Lag 0 (Same day [0-24hrs])

Eight studies with eleven outcomes (n = 1,019,062) were reported for OR. Kuzma et al. [26] reported a reduced risk of STEMI with PM_10_ exposure, while all other studies reported an increased risk within their respective groups/subgroups (MI, STEMI, NSTEMI). Statistical pooling was appropriate due to statistical homogeneity (I^2^ = 60%) (Figure 4). There was an overall increased risk of MI (RR = 1.01, 95% CI: 1.00-1.02), and it was statistically significant (P-value = 0.01). Evidence quality was rated high on the GRADE scale (S8 Table).

**Figure 4.**
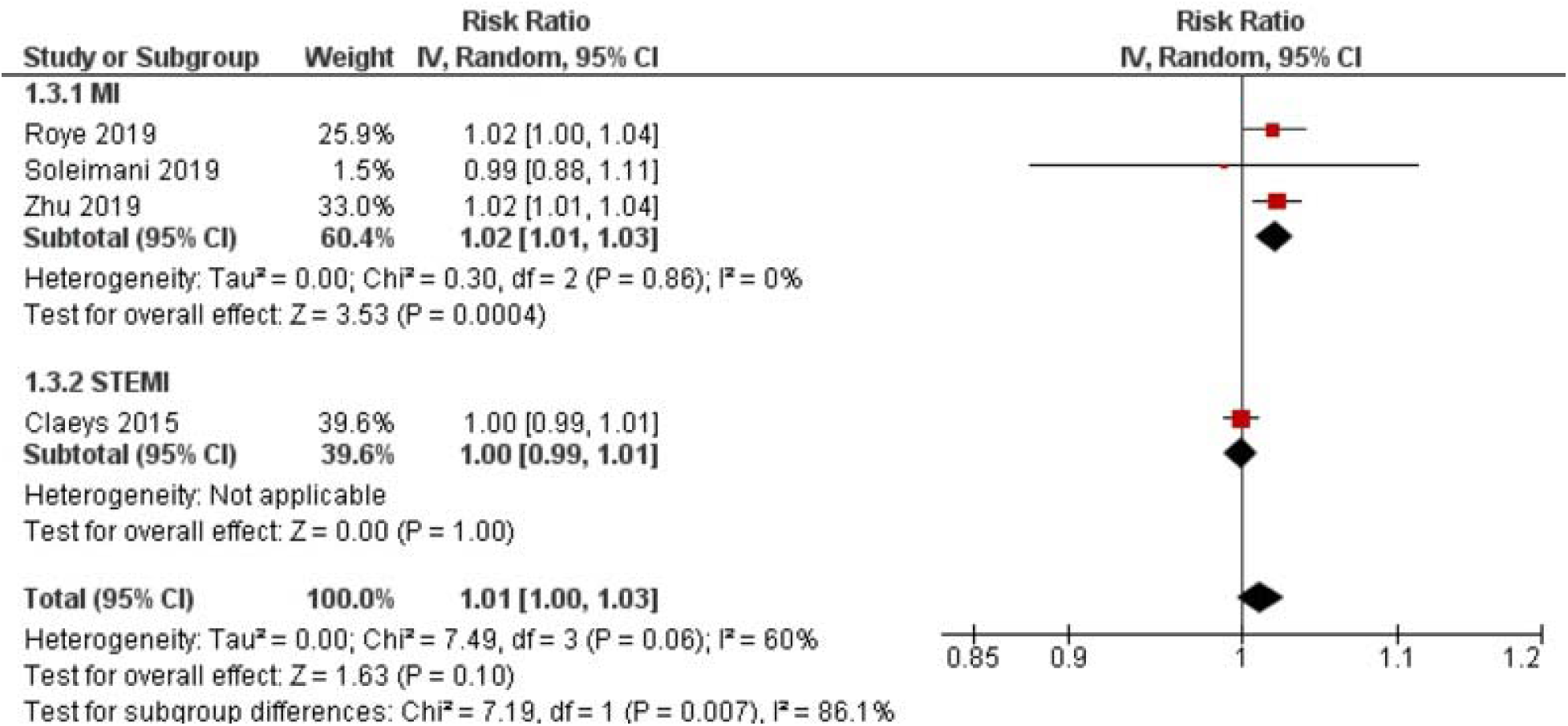
Lag 2 RR for MI after 10 μg/m^3^ increase in PM_10_ exposure.

#### 3.4.5. Odds Ratio – Lag 1 (1-3 days [24-96hrs])

Seven studies with ten outcomes (n = 1,007,634) were reported for OR. Kuzma et al. [26] reported a reduced risk of STEMI with PM_10_ exposure, while all other studies reported an increased risk within their respective groups/subgroups (MI, STEMI, NSTEMI). Statistical pooling was appropriate due to statistical homogeneity (I^2^ = 50%) (Figure 5). There was an overall increased risk of MI (RR = 1.01, 95% CI: 1.00-1.02), and it was statistically significant (P-value = 0.02). Evidence quality was rated high on the GRADE scale (S9 Table).

**Figure 5.**
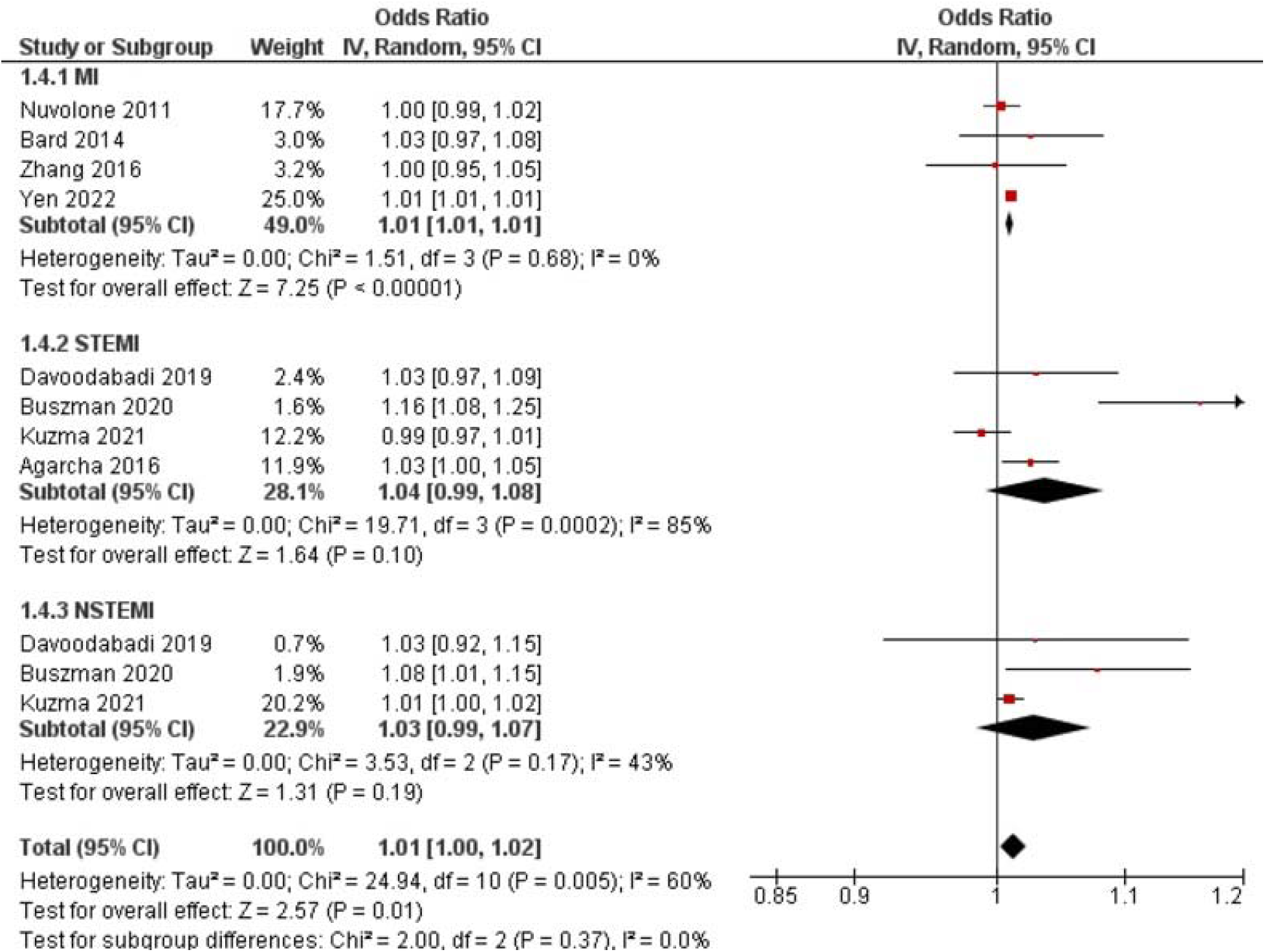
Lag 0 OR for MI after 10 μg/m^3^ increase in PM_10_ exposure.

#### 3.4.6. Odds Ratio – Lag 2 (3+ days [96hrs+])

Five studies with seven outcomes (n =27,109) were reported for OR. Zhang et al. [35] reported a reduced risk of MI with PM_10_ exposure, while all other studies reported an increased risk within their respective group/subgroup (MI, STEMI, NSTEMI). Statistical pooling was appropriate due to statistical homogeneity (I^2^ = 41%) (Figure 6). There was an overall increased risk of MI (RR = 1.01, 95% CI: 0.99-1.02), but it was not statistically significant (P-value = 0.28). Evidence quality was rated high on the GRADE scale (S10 Table).

**Figure 6.**
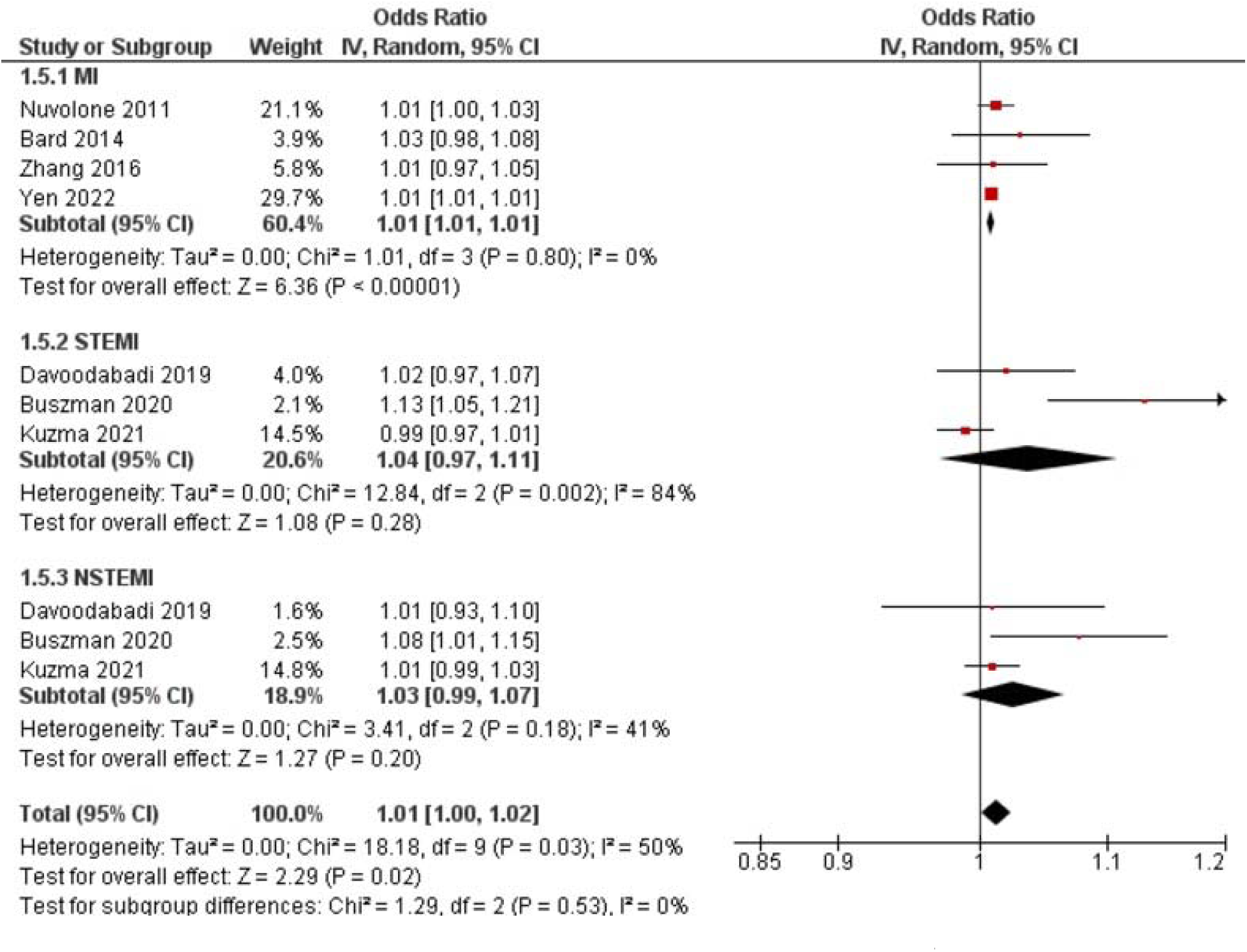
Lag 1 OR for MI after 10 μg/m^3^ increase in PM_10_ exposure.

**Figure 7.**
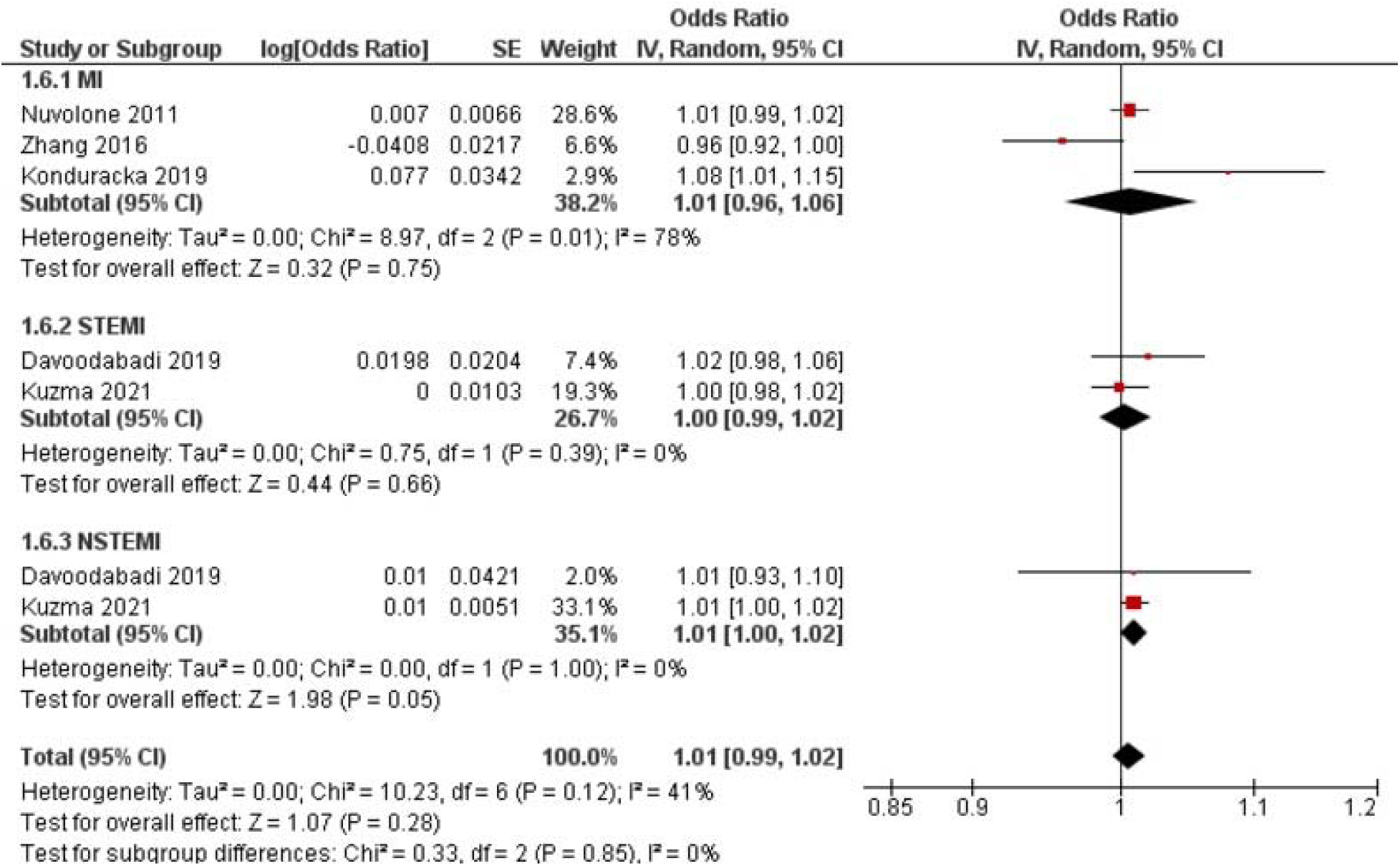
Lag 2 OR for MI after 10 μg/m^3^ increase in PM_10_ exposure.

#### 3.4.8. Hazard Ratio – Lag 2 (3+ days [96hrs+])

Six studies with six outcomes (n = 4,913,615) were reported for HR. Nuvolone et al. [28] and Konduracka et al. [25] reported an increased risk of MI with PM_10_ exposure, while Zhang et al. [35] reported a reduced risk. Statistical pooling was appropriate due to statistical homogeneity (I^2^ = 6%) (Figure 8). There was no impact on risk of MI (RR = 1.00, 95% CI: 0.99-1.01), and the result was not statistically significant (P-value = 0.69). Evidence quality was rated high on the GRADE scale (S13 Table).

**Figure 8.**
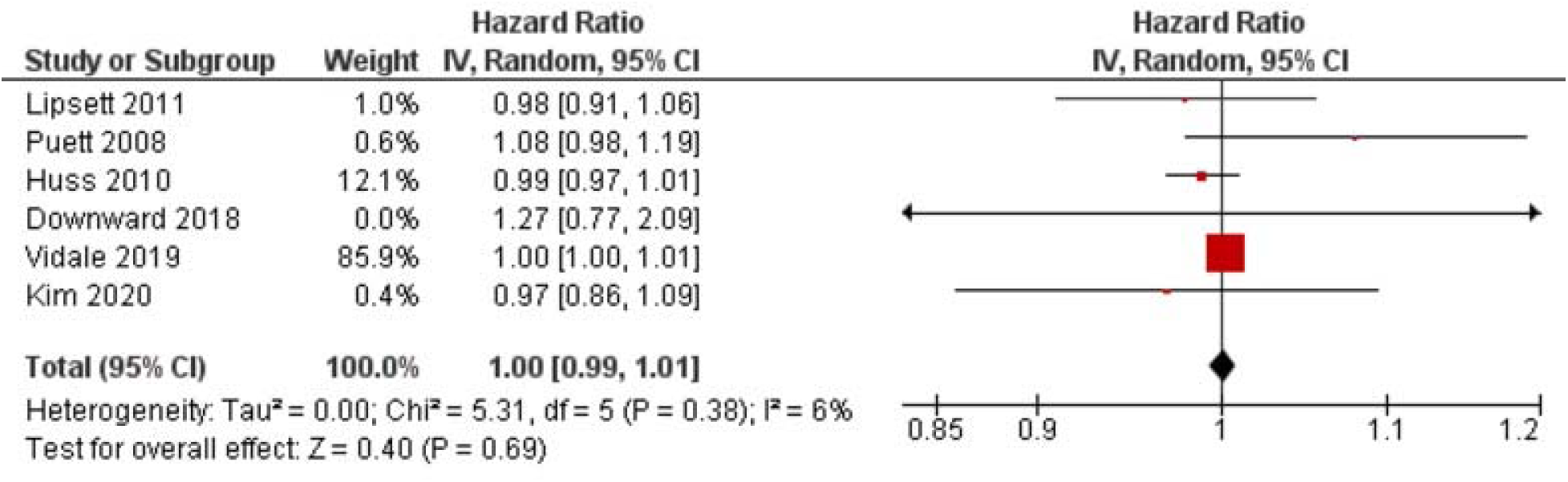
Lag 2 HR for MI after 10 μg/m^3^ increase in PM_10_ exposure.

#### 3.4.10. Sensitivity Analyses

Two sensitivity analyses were performed. The first sensitivity analysis was the exclusion of risk outcome values which had more than a 7-day post-exposure delay. This was done in order to eliminate values which had relatively long post-exposure delays lasting multiple weeks, months, or years. This sensitivity was conducted to all lag 2 forest plots and consisted of six excluded values: 1 for RR [30] and 5 for HR [22, 23, 24, 27, 29]. However, no significant differences were found in this sensitivity analysis.

The second sensitivity analysis was the exclusion of Buszman et al. [17] from the OR forest plots for lag 0 and lag 1. This was done in order to evaluate whether this study was a detrimental outlier and played a significant role in the overall risk outcome. However, no significant differences were found in this sensitivity analysis.

## 4. Discussion

This study aimed to evaluate the risk of MI after exposure to PM_10_ considering that small proportions of PM_10_ can pass the alveolar barrier, enter the blood, and contribute to cardiovascular adversities as aforementioned [5]. This was important considering that CV disease is the leading cause of death in the world [55]. Assessing potential exacerbators is crucial to reducing the burden of disease. Notably, our analysis showed an increased risk (OR=1.01; 95% CI:1.00 - 1.02) of myocardial infarction with a 10 μg/m^3^ increase in PM_10_ after a lag 0 and lag 1 delay. Considering the high prevalence of MI, the statistically significant risk increase of 1% indicates the absolute risk of PM10 is high.

Farhadi et al. [6] investigated short-term exposure to PM_2.5_ and its effects on the risk of MI. Our study and their study both looked at RR and OR as a risk outcome, and assessed studies with a case-crossover and time-series design. However, our study additionally looked at HR as a risk outcome, and assessed studies with a cohort design. Both studies also looked at the effects of lags, but lag definitions were different. Our study looked at lags with short delays (lag 0 = 0-24 hours, lag 1 = 24-96 hours, lag 2 = >96 hours), whereas Farhadi et al. [6] looked at long delays (short follow-up period = <4 years, long follow-up period = >4 years). The results of their study are comparable to our findings as the RR and OR in our meta-analysis were similar. Their meta-analysis reported a RR of 1.02 (95% CI: 1.01–1.03; P-value ≤ 0.0001) [6]. However, our results had more homogeneity, likely due to stratification of studies by risk outcome measures and smaller lag ranges. Our statistical analysis yielded ORs of 1.01 (95% CI 1.00–1.02; P-value <0.05) for lag 0 and lag 1. These results indicate that while the risk of MI from exposure to PM_10_ is 1% less in comparison to PM_2.5_ exposure, there is a potential impact of PM_2.5-10_ pollutants on MI risk. This indicates that the risk of PM_2.5-10_ entering the bloodstream through the alveolar barrier and causing CV adversities, is similar to that of PM_2.5_.

When assessing CV health concerns related to particulate matter, PM_10_ should not be discredited – especially in communities of low socioeconomic status, as these populations are most vulnerable to the health effects of air pollution [56].

### 4.1. Strengths and Limitations

This study had some limitations. The first one was the varying adjustment models utilized by included studies. There are multiple participant confounders (hypertension, diet, sleep, etc.) to consider since PM_10_ association to MI risk is observational, which could explain potential differences in MI risk outcomes. The retrospective nature of case-crossover and time-series designs is also a potential drawback considering the ambiguity of lag definitions. Another factor to consider is the heterogeneity of the studies. All studies varied in study design, population and age characteristics, and the exposure assessment methods. This review’s studies included a variety of population sizes from various places, which can limit how the results can be applied to specific people or environments. The strength and direction of the relationship between PM_10_ exposure and the risk of MI may be influenced by variables in air pollution sources, climate, and demographic factors.

### 4.2. Next Steps

To enhance the precision and applicability of the findings, certain measures can be taken in the future. The compatibility and consistency of studies assessing MI risk can be increased by standardizing a specific risk outcome measure. While conversion algorithms exist for RR and OR, they depend on an estimated incidence variable (P0) which can constrain data synthesis validity. Future research should seek to study the distinct risk of STEMI and NSTEMI. Understanding these MI subtypes’ diverse relationships with PM_10_ exposure can offer more specialized insights for prevention. Comprehensive adjustment models should also be more widely employed to ensure confounding variables are taken into account for all associative PM_10_ studies. This will allow for more robust and reliable risk estimates. By taking these additional actions, researchers can improve the accuracy and application of the findings, and clarify the link between exposure to PM_10_ and the risk of MI.

## 5. Conclusions

The results of this meta-analysis showed that there is an increased risk of MI with a 10μg/m^3^ increase in PM_10_ exposure (OR=1.01; 95% CI:1.00 - 1.02). Risk of MI is also marginal between PM_10_ and PM_2.5_ exposure. It is important to recognize and assess the CV impacts of PM_10_ alongside other pollutants. This can help guide environmental policy, individual-level preventive measures, and global public health initiatives focused on lessening the disease burden of MI caused by PM_10_ exposure.

## Supporting information

PRISMA Checklist

## Data Availability

All data is available in the manuscript.

## 6. Acknowledgments

We have no funding source or competing interests to report. All data used within this review can be found in the manuscript and supporting information.

## 8. Supporting Information Captions

**S1 Table. Search strategy**

**S2 Table. Adjusted confounding variables for studies included in the meta-analysis**

**S3 Table. Study characteristics of remaining studies included in the review**

**S4 Table. Methodological risk of bias assessment**

**S5 Table. GRADE assessment for RR – Lag 1 (1-3 days [24-96hrs]).** O = Criteria Satisfied, -1 = Criteria Unsatisfied

**S6 Table. GRADE assessment for RR – Lag 1 (1-3 days [24-96hrs]).** O = Criteria Satisfied, -1 = Criteria Unsatisfied

**S7 Table. GRADE assessment for RR – Lag 2 (3+ days [96hrs+]).** O = Criteria Satisfied, -1 = Criteria Unsatisfied

**S8 Table. GRADE assessment for OR – Lag 0 (Same day [0-24hrs]).** O = Criteria Satisfied, - 1 = Criteria Unsatisfied

**S9 Table. GRADE assessment for OR – Lag 1 (1-3 days [24-96hrs]).** O = Criteria Satisfied, -1 = Criteria Unsatisfied

**S10 Table. GRADE assessment for OR – Lag 2 (3+ days [96hrs+]).** O = Criteria Satisfied, -1 = Criteria Unsatisfied

**S11 Table. GRADE assessment for HR – Lag 0 (Same day [0-24hrs]).** O = Criteria Satisfied, -1 = Criteria Unsatisfied

**S12 Table. GRADE assessment for HR – Lag 1 (1-3 days [24-96hrs]).** O = Criteria Satisfied, - 1 = Criteria Unsatisfied

**S13 Table. GRADE assessment for HR – Lag 2 (3+ days [96hrs+]).** O = Criteria Satisfied, -1 = Criteria Unsatisfied

**S1 Figure. Lag 0 HR for MI after 10** μ**g/m^3^ increase in PM_10_ exposure.**

**S2 Figure. Lag 1 HR for MI after 10** μ**g/m^3^ increase in PM_10_ exposure.**

**S3 Figure. Protocol**

