## Supplemental Table 1 for "Association between PM_10_ exposure and risk of myocardial infarction in adults: a systematic review and meta-analysis"

| # | Search Terms | OVID Medline | Embase | CINAHL | Web of Science |
|---|---|---|---|---|---|
| 1 | “Air pollution” | 71,529 | 102,169 | 12,364 | N/A |
| 2 | “Particulate matter” | 39,216 | 72,632 | 5,438 | N/A |
| 3 | “PM10” | 8,899 | 14,177 | 1,009 | N/A |
| 4 | 1 or 2 or 3 | 92,580 | 147,058 | 14,492 | 214,878 |
| 5 | “Myocardial infarction” | 277,001 | 477,620 | 71,936 | N/A |
| 6 | “Heart attack” | 194,326 | 432,356 | 73,945 | N/A |
| 7 | 5 or 6 | 279,848 | 479,862 | 73,945 | 335,893 |
| 8 | “Adult” | 8,383,747 | 10,724,393 | 2,210,812 | 1,902,365 |
| 9 | 4 and 7 and 8 | 286 | 533 | 125 | 155 |
