## Supplemental Table 2 for "Association between PM_10_ exposure and risk of myocardial infarction in adults: a systematic review and meta-analysis"

| # | Author/ publication year | Adjusted confounding variables |
|---|---|---|
| 1 | Argacha 2016 | Age, comorbidities, smoking status, ambient air temperature, day of the week and season |
| 2 | Bard 2014 | Holidays, meteorological variables (daily maximum temperature, maximum atmospheric pressure, and mean relative humidity), and influenza epidemics |
| 3 | Bhaskaran 2011 | Daily mean temperature, daily relative humidity, daily levels of influenza and respiratory syncytial virus, day of week, and holidays |
| 4 | Buszman 2020 | Unadjusted |
| 5 | Cheng 2021 | Temperature, relative humidity, public holidays, circulating levels of influenza and respiratory syncytial virus |
| 6 | Claeys 2015 | Unadjusted |
| 7 | Collart 2017 | Seasonality, long-term trend, day of the week, and temperature |
| 8 | Davoodabadi 2019 | Temperature, dew point, and wind speed |
| 9 | Downward 2018 | Smoking status (including number of cigarettes and duration of smoking), diet (intake of fruit and vegetables), alcohol consumption, BMI, recruitment year, gender, marital status, education level, and area-level economic status |
| 10 | Huss 2010 | Sex, civil status (single, married, divorced, widowed), nationality (Swiss, other), educational level (primary, secondary, tertiary), setting (urban, rural), language region (German, French, Italian), type of building (older than 30 years without renovation versus other), socioeconomic status of the municipality, noise, and distance.<br>Analysis restricted to persons who lived at least 15 years at the same place of residence. |
| 11 | Kim 2020 | PM2.5, sex, age, income, smoking, alcohol, obese, physical activity, comorbidity for hypertension, diabetes, hyperlipidemia, area-level gross regional domestic products, % of high school graduated or more, % of elderly |
| 12 | Konduracka 2019 | Temperature changes, relative humidity, and atmospheric pressure, infections prior to hospital admission |
| 13 | Kuzma 2021 | Day of the week, public holidays, daily relative humidity, atmospheric pressure, and temperature |
| 14 | Lipsett 2011 | Age, race, smoking status, total pack-years, body mass index, marital status, alcohol consumption, second-hand smoke exposure at home, dietary fat, dietary fiber, dietary calories, physical activity, menopausal status, hormone therapy use, family history of MI or stroke, blood pressure medication, and aspirin use, and for contextual variables (income, income inequality, education, population size, racial composition, and unemployment) |
| 15 | Nuvolone 2011 | Age, gender, body mass index, influenza epidemics, population decreases during |

| | | |
|---|---|---|
| | | vacation periods, holidays, and apparent temperature |
| 16 | Puett 2008 | Age in months, adjusting for state of residence, year, and season |
| 17 | Roye 2019 | Influenza cases |
| 18 | Soleimani 2019 | Hypertension, high blood pressure, relative humidity |
| 19 | Vidale 2017 | Age, race, smoking status, total pack-years, body mass index, marital status, alcohol consumption, second-hand smoke exposure at home, dietary fat, dietary fiber, dietary calories, physical activity, menopausal status, hormone therapy use, family history of MI or stroke, blood pressure medication, and aspirin use, and for contextual variables (income, income inequality, education, population size, racial composition, and unemployment). |
| 20 | Yang 2022 | Temperature, relative humidity, time, day of the week, and holiday |
| 21 | Yen 2022 | Included gender, age group, low-income household, major injury card, Charlson comorbidity index, and meteorological data (daily temperature and relative humidity). |
| 22 | Zhang 2016 | Temperature and relative humidity |
| 23 | Zhu 2019 | Public holidays (Holiday) and day of week (DOW) |
