## Supplemental Table 3 for "Association between PM_10_ exposure and risk of myocardial infarction in adults: a systematic review and meta-analysis"

| # | Author/publication year | Study Design | Country | Sample size (n) | # Male (%) | Risk Measure | Exposure Increment | Lag Intervals |
|---|---|---|---|---|---|---|---|---|
| 1 | Akbarzadeh 2018 [37] | Case-crossover | Iran | 208 | 182 (88.3) | RR | 3.61μg/$m^3$ | Lag 0: 24hrs post-exposure<br>Lag 1: Average of 3 weeks post-exposure |
| 2 | Atkinson 2013 [38] | Cohort | United Kingdom | 13,956 | 8,471 (60.7) | HR | IQR change: 3μg/$m^3$ | Lag 1: 5-year delay |
| 3 | Cheng 2009 [39] | Case-crossover | Taiwan | 9,349 | NR | OR | IQR change: 61.94μg/$m^3$ | Lag 0-2: Average of same day and previous 2 days |
| 4 | Cramer 2020 [40] | Time-series | Denmark | 22,882 | 0 (0) | HR | IQR change: 5.5μg/$m^3$ | Lag 1: 1-year mean<br>Lag 2: 3-year mean |
| 5 | Kim 2017 [41] | Cohort | South Korea | 136,094 | 66,851 (49.1) | HR | 1μg/$m^3$ | Lag 1: 7-year delay |
| 6 | Kuzma 2020 [42] | Case-crossover | Poland | 1,790 | 1,172 (65.5) | OR | IQR change: 15.1μg/$m^3$ | Lag 0: NR<br>Lag 1: NR |
| 7 | Lee 2017 [43] | Cohort | South Korea | 37,880 | 26,787 (70.7) | HR | By Quintile: varying increments | Lag 0: 24hrs post-exposure |
| 8 | Liu 2020 [44] | Case-crossover | Canada | 6,142 | 4,482 (73.0) | OR | IQR change: 16μg/$m^3$ | Single-day lags: Lag 0, 1, 2<br>Multiple-day lags: Lag 0–3, 0–5 |
| 9 | Pan 2019 [45] | Case-crossover | Taiwan | 898 | 745 (83.0) | OR | IQR change: 50μg/$m^3$ | Single-day lags: Lag 0, 1, 2, 3 |
| 10 | Rasche 2018 [46] | Case-crossover | Germany | 693 | 466 (67.2) | OR | 10 ug/$m^3$ | Single-day lags: Lag 1, 2, 3 |
| 11 | Rodins 2020 [47] | Cohort | Germany | 4,105 | 1,950 (47.5) | HR | 1 ug/$m^3$ | Lag 1: 14-year delay |
| 12 | Sahlen 2019 [48] | Case-crossover | Sweden | 14,601 | NR | OR | IQR change: 26.5μg/$m^3$ | Lag 1-24: Hourly delays per lag |

| | | | | | | | | |
|---|---|---|---|---|---|---|---|---|
| 13 | Sen 2016 [49] | Case-crossover | Turkey | 402 | 310 (77.1) | RR | 5 ug/m$^3$ | Single-day lags: Lag 0, 1, 2, 7 |
| 14 | Wang 2016 [50] | Case-crossover | China | 972 | 515 (53.0) | OR | 50 ug/m$^3$ | Lag 0: Same day |
| 15 | Wichmann 2013 [51] | Case-crossover | Sweden | 28,215 | 16,627 (58.9) | OR | IQR change: 10.4μg/m$^3$ | Lag 0: Same day<br>Lag 1: 1-day delay<br>Lag 0-1: Average of same day and 1 day previous |
| 16 | Wichmann 2014 [52] | Case-crossover | Sweden | 28,215 | 16,627 (58.9) | OR | IQR change: 10.4μg/m$^3$ | Lag 0: Same day<br>Lag 1: 1-day delay<br>Lag 0-1: Average of same day and 1 day previous |
| 17 | Wolf 2015 [53] | Cohort | Germany | 15,417 | 11,378 (73.8) | RR | 24.3 ug/m$^3$ | Lag 0: Same day<br>Lag 1: 1-day delay<br>Lag 0-5: Average of same day and previous 5 days |
| 18 | Yu 2018 [54] | Case-crossover | China | 5,545 | 3,492 (63.0 | RR | 10 ug/m$^3$ | Single-day lags: Lag 0, 1, 2, 3, 4, 5, 6<br>Multiple-day lags: Lag 0–1, 0–2, 0–3, 0–4, 0–5, 0–6 |
