## Supplemental Table 4 for "Association between PM_10_ exposure and risk of myocardial infarction in adults: a systematic review and meta-analysis"

| ID# | Study | Q1 | Q2 | Q3 | Q4 | Q5 | Q6 | Q7 | Q8 | Q9 | Q10 | Q11 | Q12 | Q13 | Q14 | Score |
|---|---|---|---|---|---|---|---|---|---|---|---|---|---|---|---|---|
| **#226** | Akbarzadeh 2018 | O | O | O | O | O | O | O | O | O | O | O | NA | O | O | Good |
| **#298** | Argacha 2016 | O | O | O | O | O | O | O | O | O | O | O | NA | O | O | Good |
| **#379** | Atkinson 2013 | O | O | O | O | O | O | O | O | O | O | O | NA | O | O | Good |
| **#355** | Bard 2014 | O | O | O | O | O | O | O | O | O | O | O | NA | O | O | Good |
| **#415** | Bhaskaran 2011 | O | O | O | O | O | O | O | O | O | O | O | NA | O | O | Good |
| **#129** | Buszman 2020 | O | O | O | O | O | O | O | O | O | O | O | NA | O | – | Fair |
| **#89** | Cheng 2021 | O | O | O | O | O | O | O | O | O | O | O | NA | O | O | Good |
| **#453** | Cheng 2009 | O | O | O | O | O | O | O | O | O | O | O | NA | O | O | Good |
| **#44** | Claeys 2015 | O | O | O | O | O | O | O | O | O | O | O | NA | O | – | Fair |
| **#288** | Collart 2017 | O | O | O | O | O | O | O | O | O | O | O | NA | O | O | Good |
| **#140** | Cramer 2020 | O | O | O | O | O | O | O | O | O | O | O | NA | O | O | Good |
| **#193** | Davoodabadi 2019 | O | O | O | O | O | O | O | O | O | O | O | NA | O | O | Good |
| **#237** | Downward 2018 | O | O | O | O | O | O | O | O | O | O | O | NA | O | O | Good |
| **#450** | Huss 2010 | O | O | O | O | O | O | O | O | O | O | O | NA | O | O | Good |
| **#263** | Kim 2017 | O | O | O | O | O | O | O | O | O | O | O | NA | O | O | Good |
| **#130** | Kim 2020 | O | O | O | O | O | O | O | O | O | O | O | NA | O | O | Good |
| **#753** | Konduracka 2019 | O | O | O | O | O | O | O | O | O | O | O | NA | O | O | Good |
| **#133** | Kuzma 2020 | O | O | O | O | O | O | O | O | O | O | O | NA | O | – | Fair |
| **#105** | Kuzma 2021 | O | O | O | O | O | O | O | O | O | O | O | NA | O | O | Good |
| **#265** | Lee 2017 | O | O | O | O | O | O | O | O | O | O | O | NA | O | O | Good |
| **#424** | Lipsett 2011 | O | O | O | O | O | O | O | O | O | O | O | NA | O | O | Good |
| **#64** | Liu 2020 | O | O | O | O | O | O | O | O | O | O | O | NA | O | O | Good |
| **#428** | Nuvolone 2011 | O | O | O | O | O | O | O | O | O | O | O | NA | O | O | Good |
| **#195** | Pan 2019 | O | O | O | O | O | O | O | O | O | O | O | NA | O | O | Good |
| **#462** | Puett 2008 | O | O | O | O | O | O | O | O | O | O | O | NA | O | O | Good |
| **#236** | Rasche 2018 | O | O | O | O | O | O | O | O | O | O | O | NA | O | O | Good |
| **#137** | Rodins 2020 | O | O | O | O | O | O | O | O | O | O | O | NA | O | O | Good |
| **#201** | Roye 2019 | O | O | O | O | O | O | O | O | O | O | O | NA | O | O | Good |
| **#758** | Sahlen 2019 | O | O | O | O | O | O | O | O | O | O | O | NA | O | O | Good |
| **#318** | Sen 2016 | O | O | O | O | O | O | O | O | O | O | O | NA | O | O | Good |
| **#183** | Soleimani 2019 | O | O | O | O | O | O | O | O | O | O | O | NA | O | O | Good |

| | | | | | | | | | | | | | | | | |
|---|---|---|---|---|---|---|---|---|---|---|---|---|---|---|---|---|
| **#262** | Vidale 2017 | O | O | O | O | O | O | O | O | O | O | O | NA | O | O | Good |
| **#312** | Wang 2016 | O | O | O | O | O | O | O | O | O | O | O | NA | O | O | Good |
| **#380** | Wichmann 2013 | O | O | O | O | O | O | O | O | O | O | O | NA | O | O | Good |
| **#367** | Wichmann 2014 | O | O | O | O | O | O | O | O | O | O | O | NA | O | O | Good |
| **#347** | Wolf 2015 | O | O | O | O | O | O | O | O | O | O | O | NA | O | O | Good |
| **#564** | Yang 2022 | O | O | O | O | O | O | O | O | O | O | O | NA | O | O | Good |
| **#9** | Yen 2022 | O | O | O | O | O | O | O | O | O | O | O | NA | O | O | Good |
| **#252** | Yu 2018 | O | O | O | O | O | O | O | O | O | O | O | NA | O | O | Good |
| **#323** | Zhang 2016 | O | O | O | O | O | O | O | O | O | O | O | NA | O | O | Good |
| **#759** | Zhu 2019 | O | O | O | O | O | O | O | O | O | O | O | NA | O | O | Good |
| O = Criteria satisfied; – = Criteria unsatisfied; NA = Not applicable | | | | | | | | | | | | | | | | |
