## Supplemental Table 6 for "Association between PM_10_ exposure and risk of myocardial infarction in adults: a systematic review and meta-analysis"

| **GRADE Criteria** | **RoB** | **Inconsistency** | **Indirectness** | **Imprecision** | **Publication Bias** | **Large magnitude of effect** | **Dose-response gradient** | **Residual Confounding** | **<u>Quality</u>** |
|---|---|---|---|---|---|---|---|---|---|
| **Result** | O | –1 | O | O | O | O | O | O | Moderate |
| **Reason** | 4 Good<br>0 Fair | $I^2$ = 72% | Already screened for | 1.01 (0.99, 1.03)<br><br>CI limits do not cross the 25th percentile. | <5 studies | NA | NA | NA | |
