## Supplemental Table 9 for "Association between PM_10_ exposure and risk of myocardial infarction in adults: a systematic review and meta-analysis"

| **GRADE Criteria** | **RoB** | **Inconsistency** | **Indirectness** | **Imprecision** | **Publication Bias** | **Large magnitude of effect** | **Dose-response gradient** | **Residual Confounding** | **<u>Quality</u>** |
|---|---|---|---|---|---|---|---|---|---|
| **Result** | O | O | O | O | O | O | O | O | High |
| **Reason** | 6 Good<br>1 Fair | $I^2$ = 50% | Already screened for | 1.01 (1.00, 1.02)<br>CI limits do not cross the 25th percentile. | 7 studies<br>10 outcomes<br>Symmetrical | NA | NA | NA | |
