## Supplemental Table 10 for "Association between PM_10_ exposure and risk of myocardial infarction in adults: a systematic review and meta-analysis"

| GRADE Criteria | RoB | Inconsistency | Indirectness | Imprecision | Publication Bias | Large magnitude of effect | Dose-response gradient | Residual Confounding | Quality |
|---|---|---|---|---|---|---|---|---|---|
| **Result** | O | O | O | O | O | O | O | O | High |
| **Reason** | 5 Good<br>0 Fair | $I^2 = 41\%$ | Already screened for | 1.01 (0.99, 1.02)<br>CI limits do not cross the 25th percentile. | 5 studies<br>7 outcomes<br>Symmetrical | NA | NA | NA | |
