## Supplemental Table 11 for "Association between PM_10_ exposure and risk of myocardial infarction in adults: a systematic review and meta-analysis"

| **GRADE Criteria** | **RoB** | **Inconsistency** | **Indirectness** | **Imprecision** | **Publication Bias** | **Large magnitude of effect** | **Dose-response gradient** | **Residual Confounding** | **Quality** |
| --- | --- | --- | --- | --- | --- | --- | --- | --- | --- |
| **Result** | NA | NA | NA | NA | NA | NA | NA | NA | NA |
| **Reason** | 1 Good<br>0 Fair | 1 study | Already screened for | 1.00 (1.00, 1.01)<br>CI limits do not cross the 25th percentile. | <5 studies | NA | NA | NA | |
