## Supplemental Figure 2 for "Association between PM_10_ exposure and risk of myocardial infarction in adults: a systematic review and meta-analysis"

| Study or Subgroup | Weight | Hazard Ratio IV, Random, 95% CI | Hazard Ratio IV, Random, 95% CI |
|---|---|---|---|
| Vidale 2019 | 100.0% | 1.00 [1.00, 1.01] | |
| **Total (95% CI)** | **100.0%** | **1.00 [1.00, 1.01]** | |

Heterogeneity: Not applicable
Test for overall effect: Z = 0.38 (P = 0.70)

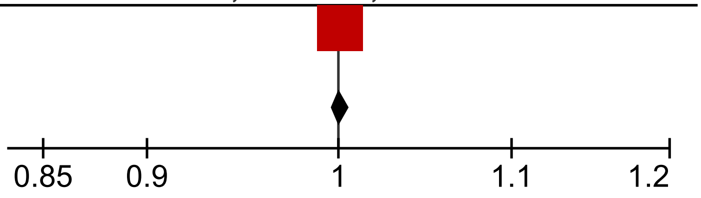
