## Supplemental Figure 3 for "Association between PM_10_ exposure and risk of myocardial infarction in adults: a systematic review and meta-analysis"

To enable PROSPERO to focus on COVID-19 submissions, this registration record has undergone basic automated checks for eligibility and is published exactly as submitted. PROSPERO has never provided peer review, and usual checking by the PROSPERO team does not endorse content. Therefore, automatically published records should be treated as any other PROSPERO registration. Further detail is provided here.

### Citation

Kleiton Strobl, Syed Irfan, Hassan Masood, Noor Latif, Om Kurmi. A systematic review and meta-analysis of PM10 exposure on risk of myocardial infarction. PROSPERO 2023 CRD42023409796 Available from: https://www.crd.york.ac.uk/prospero/display_record.php?ID=CRD42023409796

### Review question

Within adults (aged 18 or older), does high PM10 exposure result in an increased risk of myocardial infarction when compared to low PM10 exposure?

### Searches

The Ovid MEDLINE, Web of Science, and EBSCO CINAHL databases were searched on January 17, 2023, which included all studies prior to this date. There is no limitation for language.

### Types of study to be included

Inclusion criteria includes: time-series or case-crossover studies

Exclusion criteria includes: secondary studies (narrative reviews, systematic reviews, meta-analyses, scoping reviews)

### Condition or domain being studied

A particulate pollutant is a tiny or microscopic mixture of solid and liquid particles that are suspended in the air. Particulate matter may be emitted by a variety of human activities, including vehicle emissions, smoke, dust, and ash from industrial processes. There are three primary categories of particulate matter: coarse particles (PM10), small particles (PM2.5), and ultrafine particles (PM0.1). Generally speaking, the origins and health consequences of these particle sizes vary.

Short-term health consequences from exposure to small particles include irritation of the eyes, nose, throat, and lungs, as well as coughing, sneezing, runny nose, and shortness of breath. In addition to impairing lung function, exposure to tiny particles can exacerbate existing medical disorders, including asthma and heart disease. Research has also shown that exposure to particulate matter can also elevate the risk of cardiovascular events. However, particulate matter 10 exposure has been poorly studied in relation to myocardial infarction in adults. As such, this study aims to review and meta-analyze this association, specifically looking at risk of myocardial infarction in adults.

### Participants/population

Inclusion criteria includes: population ≥ 18 years of age, and participants have not had MI previously.

Exclusion criteria includes: population < 18 years of age (children), and participants that have had MI previously.

### Intervention(s), exposure(s)

Inclusion criteria includes: PM10 as an exposure

Exclusion criteria includes: studies which only look at PM2.5 as an exposure

### Comparator(s)/control

Our control includes populations with low exposure to PM10.

### Main outcome(s)

The main outcome is the risk of myocardial infarction (relative risk (RR), odds ratio (OR), prevalence).

### Additional outcome(s)

None

### Data extraction (selection and coding)

Two individuals screened the abstracts of all studies from the literature search by following the predefined inclusion and exclusion criteria. Two separate researchers resolved conflicts of this screening process to reach consensus of studies to include for further analysis.

### Risk of bias (quality) assessment

Publication bias will be determined by assessing funnel plot symmetricity, and performing a Begg's and Egger's test. Statistical heterogeneity (inconsistency) will be assessed by analyzing the $I^2$ statistic.

### Strategy for data synthesis

Study attributes will be provided in tables according to PRISMA guidelines. Meta-analysis will be performed in Review Manager 5.4.1. We will use the random effects analysis model to measure heterogeneity amongst all screened studies.

### Analysis of subgroups or subsets

If sufficient data exists, stratification according to age will be undertaken.

### Contact details for further information

Kleiton Strobl

### Organisational affiliation of the review

McMaster University

### Review team members and their organisational affiliations

Kleiton Strobl. McMaster University

Syed Irfan. McMaster University

Hassan Masood. McMaster University

Noor Latif. McMaster University

Dr Om Kurmi. McMaster University

### Type and method of review

Meta-analysis, Systematic review

### Anticipated or actual start date

17 January 2023

### Anticipated completion date

30 April 2023

### Funding sources/sponsors

None

### Conflicts of interest

None known

### Language

English

### Country

Canada

### Stage of review

Review Ongoing

### Subject index terms status

Subject indexing assigned by CRD

### Subject index terms

Adult; Air Pollution; Humans; Myocardial Infarction; Particulate Matter

### Date of registration in PROSPERO

31 March 2023

### Date of first submission

21 March 2023

### Stage of review at time of this submission

| Stage | Started | Completed |
|---|---|---|
| Preliminary searches | Yes | Yes |
| Piloting of the study selection process | Yes | Yes |
| Formal screening of search results against eligibility criteria | Yes | No |
| Data extraction | No | No |
| Risk of bias (quality) assessment | No | No |
| Data analysis | No | No |

*The record owner confirms that the information they have supplied for this submission is accurate and complete and they understand that deliberate provision of inaccurate information or omission of data may be construed as scientific misconduct.*

*The record owner confirms that they will update the status of the review when it is completed and will add publication details in due course.*

### Versions

31 March 2023

31 March 2023
